# Policies on Artificial Intelligence Chatbots Among Academic Publishers: A Cross-Sectional Audit

**DOI:** 10.1101/2024.06.19.24309148

**Authors:** Daivat Bhavsar, Laura Duffy, Hamin Jo, Cynthia Lokker, R. Brian Haynes, Alfonso Iorio, Ana Marusic, Jeremy Y. Ng

## Abstract

**Background:** Artificial intelligence (AI) chatbots are novel computer programs that can generate text or content in a natural language format. Academic publishers are adapting to the transformative role of AI chatbots in producing or facilitating scientific research. This study aimed to examine the policies established by scientific, technical, and medical academic publishers for defining and regulating the responsible authors’ use of AI chatbots.

**Methods:** This study performed a cross-sectional audit on the publicly available policies of 163 academic publishers, indexed as members of the International Association of the Scientific, Technical, and Medical Publishers (STM). Data extraction of publicly available policies on the webpages of all STM academic publishers was performed independently in duplicate with content analysis reviewed by a third contributor (September 2023 - December 2023). Data was categorized into policy elements, such as ‘proofreading’ and ‘image generation’. Counts and percentages of ‘yes’ (i.e., permitted), ‘no’, and ‘N/A’ were established for each policy element.

**Results:** A total of 56/163 (34.4%) STM academic publishers had a publicly available policy guiding the authors’ use of AI chatbots. No policy allowed authorship accreditations for AI chatbots (or other generative technology). Most (49/56 or 87.5%) required specific disclosure of AI chatbot use. Four policies/publishers placed a complete ban on the use of AI tools by authors.

**Conclusions:** Only a third of STM academic publishers had publicly available policies as of December 2023. A re-examination of all STM members in 12-18 months may uncover evolving approaches toward AI chatbot use with more academic publishers having a policy.

## Introduction

Over 350 years ago, the first academic journals were published with the purpose of disseminating research findings in a widely available and structured manner, to increase scientific knowledge [1]. Today, academic publishers and their journals have become the main source of scientific findings and means of communication, offering quick and convenient access to information [1]. Nearly 1.5 million scholarly articles are published each year and are accessed by approximately 10-15 million readers [2, 3]. The number of publications has been increasing exponentially and being so widely accessible online, more people than ever before read scholarly articles [3].

The journal publishing process involves multiple individuals—authors, peer-reviewers, editors, and academic publishers. These groups are responsible for ensuring that the content published is correct and relevant to their respective field. Manuscripts submitted by authors are peer reviewed by experts in the field. Once accepted, the manuscript is published and supports dissemination of research findings [3]. To promote publication integrity, publishers provide guidelines and construct policies for authors, peer-reviewers, and editors to follow [3]. Publication integrity is the adherence to ethical and professional practices while maintaining honesty and transparency across all aspects of research [4]. While policies surrounding topics such as copyright and ethics have long existed in academic publication, the creation of new policies to parallel societal and technological advances is an ongoing process essential to maintaining publication integrity [1]. Technological advances, such as artificial intelligence (AI), are on the rise and continue to become more accessible, raising the concern of the growing dependence on AI tools in the process of academic publication.

AI is an interdisciplinary field of science and technology that plays an expanding, dominating role in everyday human lives. AI refers to the design of machines that can mimic or surpass the mental capacities and intelligence of humans [5, 6]. AI developments since the 1950s have led to advances that allow computers and computer-controlled robots to analyze, generalize, problem-solve, and adapt to new circumstances [7]. The ability to self-learn—a crucial component of AI—coupled with the recent advances in computer science over the past two decades, has allowed AI to become increasingly integrated into various industries with tremendous potential. An important advancement in AI technology is the introduction of AI chatbots, which are easily accessible tools on the internet that can rapidly generate user-prompted outputs [8].

Over the past decade, AI chatbots have become increasingly powerful and practical tools for various fields, particularly education, healthcare, and research [9]. AI chatbots, such as ChatGPT, imitate human conversation to provide direct, succinct responses to user prompts about various topics, including marketing and healthcare [10, 11]. For example, although ChatGPT cannot provide ‘personalized’ medical advice, AI chatbots are used by patients to provide diagnostic insights and suggest therapeutic interventions [11]. Yet, this transformative technology can go beyond conversation by producing entire manuscripts, such as reports, school essays, and scientific articles [12]. AI chatbots may be used to draft literature reviews and introduction sections of manuscripts, design experimental protocols, and perform data analysis [12]. While AI chatbots offer plenty of opportunities to support and optimize the research process, there are pertinent challenges that may arise as AI chatbot use and technology advances. For example, AI chatbots gather information from the internet to form outputs that, although convincing, may not be entirely correct [13]. For instance, when prompted by users to provide a list of sources, ChatGPT has been found on occasion to falsify citations, producing references with incorrect PubMed ID numbers and years of publication [13]. The production of content-unverified outputs by AI chatbots poses the threat of misinformation, and without technology that can accurately detect AI-generated text or images, the scientific integrity of published content is threatened. As AI chatbots are increasingly used and relied upon by medical researchers and academic publishers, they can influence how information can be accessed, organized, and disseminated, thereby reshaping the ever-evolving realm of medical knowledge [5]. Due to the manner in which content is generated from AI chatbots and the risk of misinformation, manuscript authors are left to question the ethics and policies surrounding AI use. Consequently, some academic publishers have created policies to guide the use of AI chatbots, such as ChatGPT, for authors conducting research and writing their manuscripts [14].

This study is a cross-sectional audit of policies implemented by scientific, technical, and medical publishers on the use of generative AI tools by manuscript authors. Recent work has shown that AI authorship policies are primarily set by the academic publisher, rather than individual journals [1]. Therefore, the objective is to gauge the different approaches taken by academic publishers toward the authors’ use of AI chatbots by reviewing the various elements in the current policies. This will allow us to understand the ways in which academic publishers regulate authors’ use of AI chatbots to maintain the scientific integrity of the content published in their journals and to set a baseline as chatbots become increasingly available. Policies guiding AI use by other groups, such as peer-reviewers and editors, will be considered as beyond the scope of this study.

## Methods

### Open Science Statement

The protocol for this study was registered on the Open Science Framework (OSF): <u>10.17605/OSF.IO/937ES</u>. All study materials and data are also available on the OSF: <u>10.17605/OSF.IO/6HP9R</u>.

### Approach

A cross-sectional audit was performed to examine the different types of policies that exist among academic publishers on the authors’ use of generative AI tools between the dates of September 1, 2023, and December 31, 2023. Selection criteria were established to include academic publishers with an international presence and journals across multiple disciplines of science, technology, and medicine. Relevant information, such as the permission for authors to use AI tools, was then analyzed to identify the commonalities and variations of academic publisher approaches through their policies.

### Publisher Selection

The members of the International Association of Scientific, Technical and Medical Publishers (STM) were chosen for examination of their publicly available web page policies on the authors’ use of AI chatbots. The STM is a global trade association with 163 members as of December 2023, consisting of some of the largest, most influential academic publishers according to the SCImago indexing factor, such as *Elsevier* and Springer Nature [15]. These 163 members collectively publish approximately two-thirds (66%) of all scientific journal articles for a global audience [15]. This selection criteria allowed for the assessment of the leading academic publishers’ stance on the use of AI chatbots by authors in the scholarly communication process as these approaches would reflect broader industry practices. The STM member page contains the URLs for the academic publisher home page, which were verified by Google searches [15].

### Data Extraction and Management

Examination of academic publisher policies was systematically conducted through an extensive review of their websites. Upon accessing the website, we searched for the publisher characteristics (e.g., country of publisher, date established, publisher URL) to note their background and experience. The availability of a policy was determined by examining the author guidelines and separate web pages of the academic publisher. To access these web pages, the Google search bar was used to input specific search prompts, such as ‘AI policy of [publisher name]’. If guidelines for the authors’ use of AI chatbot were left to the discretion of the journal(s) (i.e., the journal exclusively established the policy), rather than the academic publisher, then the academic publisher was considered to not have a policy. Policies for other groups, such as peer-reviewers, were excluded.

Data elements for analysis were determined by first examining the policies of five varying sized academic publishers: *Elsevier*, *JAMA Network*, *MDPI*, *Taylor & Francis*, and the *Association for Computing Machinery*. Element-based extraction allowed a ‘funnel’ approach to organizing data, with broader aspects of the policies (e.g., complete ban on AI vs. permitting the use of AI) recorded prior to the examination of narrower aspects (e.g., AI permitted for image generation vs. prohibition of image altering). The key parameters collected for analysis included the conditions under which AI use is permitted by authors (e.g., declarations of AI usage and assistance, for which specific purposes can AI tools be used), AI authorship acknowledgment (e.g., whether AI tools can be listed as an author), integrity of reproduced materials (e.g., whether AI tools can be used for writing non-methodological sections without granting authorship; whether there is a policy on verifying the accuracy of AI-generated citations), citation practices (e.g., whether AI tools can be cited as a primary source or author), adherence to formal research methodologies (e.g., whether AI tools can be used for research design, including data collection and processing), image integrity standards (e.g., whether AI tools can be used for designing or altering images and graphics), and proofreading guidelines (e.g., whether AI tools can be used for proofreading). A Microsoft Excel sheet was used to note these key policy elements for all academic publishers that had a policy (i.e., a data extraction form, see: https://osf.io/s45hn).

To ensure the accuracy and reliability of the extracted data, extraction and organization of these elements for the academic publishers with publicly available policies was performed independently and in duplicate by two reviewers, using separate data extraction forms, and data verification and interpretation of policy elements was performed by a third reviewer. Any differences were reconciled with discussions between the reviewers, in attempts to standardize the way the policy was interpreted. Once the discrepancies were resolved, the data were compiled into the data extraction form.

### Data Analysis

Basic descriptive statistics, such as frequencies and percentages, were generated through the analysis of the qualitative data. In the data extraction form on Microsoft Excel, each cell contained a ‘Yes’, ‘No’, or ‘N/A’ (‘Not Available’) for the specific policy element of each academic publisher (e.g., ‘Yes’ for the ‘Proofreading’ sub-section of the *Elsevier* policy). Additional details were provided in the cell as well, as appropriate (e.g., image altering permitted, but not image generation).

## Results

The search and extraction of publicly available policies was performed between September 1, 2023, and December 31, 2023 from the 163 academic publishing members listed on the STM association website^15^. The website URLs, obtained from the STM member page, and the founding details for each of the 163 publishers are listed in **Table 1**. A complete copy of the data extractions can be found on OSF: https://osf.io/s45hn.

**Table 1.** Use of AI Chatbots by Authors: Policy Availability of 163 Academic Publishers Indexed Under the International Association of Scientific, Technical, and Medical Publishers (STM)

| STM Member | Website URL | Founding Year | Nation of Origin | Policy Availability |
| --- | --- | --- | --- | --- |
| AIP Publishing | https://www.aip.org/ | 1931 | USA | Yes |
| American Association for the Advancement of Science | https://www.aaas.org/ | 1848 | USA | Yes |
| American Association of Critical-Care Nurses (AACN) | https://www.aacn.org/ | 1969 | USA | Yes |
| American Chemical Society | https://www.acs.org/ | 1976 | USA | Yes |
| American College of Physicians | http://www.acponline.org/ | 1915 | USA | Yes |
| American Mathematical Society | https://www.ams.org/home/page | 1888 | USA | Yes |
| JAMA Network | https://jamanetwork.com/ | 1883 | USA | Yes |
| American Physical Society | https://www.aps.org/ | 1899 | USA | Yes |
| American Physiology Society | https://www.physiology.org/?SSO=Y | 1887 | USA | Yes |
| American Psychiatric Association | https://www.psychiatry.org/ | 1844 | USA | No |
| American Psychological Association | https://www.apa.org/ | 1892 | USA | Yes |
| American Society for Parenteral and Enteral Nutrition (ASPEN) | https://www.nutritioncare.org/ | 1975 | USA | No |
| American Society of Agronomy | https://www.agronomy.org/ | 1907 | USA | Yes |
| American Society of Civil Engineers | https://www.asce.org/ | 1852 | USA | Yes |
| American Society of Clinical Oncology | https://www.asco.org/ | 1964 | USA | Yes |
| American Society of Mechanical Engineers (ASME) | https://www.asme.org/ | 1880 | USA | Yes |
| Anadem Publishing | https://anadem.com/ | N/A | USA | No |
| **Apple Academic Press Inc.** | https://www.appleacademicpress.com/ | 2008 | USA | No |
| **Aries System** | https://www.ariessys.com/ | 1986 | USA | No |
| **Association for Computing Machinery** | https://www.acm.org/ | 1947 | USA | Yes |
| **Association of American Publishers** | https://publishers.org/ | 1945 | USA | No |
| **Association of American University Presses** | https://aupresses.org/ | 1937 | USA | No |
| **Association of Learned and Professional Society Publishers** | https://www.alpsp.org/ | 1972 | USA | No |
| **Association of Medical Illustrators** | https://www.ami.org/ | 1945 | USA | No |
| **Atypon** | https://www.atypon.com/ | 1996 | USA | No |
| **BCS** | https://www.bcs.org/ | 1957 | England | No |
| **Begell House** | https://www.begellhouse.com/ | 1991 | USA | No |
| **Beilstein-Institut** | https://www.beilstein-institut.de/en/ | 1951 | Germany | No |
| **Berlin Institute for Scholarly Publishing (BISP)** | https://berlinstitute.org/ | 2020 | Germany | No |
| **BioExcel Publishing** | https://www.bioexcelpublishing.com/ | 2005 | England | No |
| **Bioscientifica** | https://www.bioscientifica.com/publishing/ | 1996 | England | No |
| **BMJ** | https://www.bmj.com/ | 1840 | England | Yes |
| **Börsenverein des Deutschen Buchhandels** | https://www.boersenverein.de/ | 1825 | Germany | No |
| **Brill** | https://brill.com/ | 1683 | Netherlands | Yes |
| **British Small Animal Veterinary Association** | https://www.bsava.com/ | 1957 | England | No |
| **British Society for Rheumatology** | https://www.rheumatology.org.uk/ | N/A | England | No |
| **Burleigh Dodds Science Publishing** | https://www.bdspublishing.com/ | 2015 | England | No |
| **CABI** | https://www.cabi.org/ | 1910 | England | Yes |
| **Cairn.info** | https://www.cairn.info/ | 2005 | France | No |
| **Cambridge Media** | https://www.cambridge-media.com.au/ | N/A | Australia | No |
| **Cambridge University Press** | https://www.cambridge.org/ | 1534 | England | Yes |
| **Canadian Science Publishing** | https://cdnsiencepub.com/ | 1929 | Canada | Yes |
| **Cardiotext Publishing** | https://cardiotextpublishing.com/ | 2008 | USA | No |
| **Charlesworth Group** | https://charlesworth-group.com/ | 1928 | England | No |
| **ChemTec Publishing** | https://chemtec.org/ | 1996 | Canada | No |
| **CHORUS** | https://www.chorusaccess.org/ | 2014 | USA | No |
| **Clarivate Analytics** | https://clarivate.com/ | 2016 | USA | No |
| **Clarke & Esposito** | https://www.ce-strategy.com/ | 2018 | USA | No |
| **Clinical Pocket Reference Ltd** | https://www.clinical-pocketreference.com/ | 2002 | England | No |
| **Compuscript** | https://compuscript.com/ | 1991 | Ireland | No |
| **Copyright Clearance Center** | https://www.copyright.com/ | 1978 | USA | No |
| **Crossref** | https://www.crossref.org/ | 2000 | USA | No |
| **CSIRO Publishing** | https://www.publish.csiro.au/ | 1995 | Australia | Yes |
| **De Gruyter** | http://www.degruyter.com/ | 1749 | Germany | Yes |
| **Delta Think Inc.** | https://deltathink.com/ | 2005 | USA | No |
| **Deutsche Ärzteverlag** | http://www.aerzteverlag.de/ | 1949 | Germany | No |
| **Digital Science** | http://www.digital-science.com/ | 2010 | England | No |
| **Dunedin Academic Press** | https://www.dunedinacademicpress.co.uk/ | 2000 | Scotland | No |
| **EB Medicine** | http://www.ebmedicine.net/ | 1999 | USA | Yes |
| **EBSCO** | https://www.ebsco.com/ | 1944 | USA | No |
| **EDP Sciences** | http://www.edpsciences.org/ | 1920 | France | Yes |
| **Elmer Press** | https://www.elmerpress.com/ | 2008 | Canada | No |
| **Elsevier** | https://beta.elsevier.com/?trial=true | 1880 | Netherlands | Yes |
| **Emerald Pressing** | http://www.emeraldpublishing.com/ | 1967 | England | Yes |
| **EMS Press** | https://ems.press/ | 1990 | Finland | No |
| **Endocrine Society** | http://www.endocrine.org/ | 1916 | USA | Yes |
| **Eurasia Academic Publishing Group** | https://eaapublishing.org/ | 2017 | China | Yes |
| **European Association for Cardiac-Thoracic Surgery** | https://www.eacts.org/ | 1986 | England | No |
| **European Respiratory Society** | http://www.ersjournals.com/ | 1990 | Switzerland | No |
| **Exon Publications** | https://exonpublications.com/ | 2020 | Australia | Yes |
| **Federation of European Publishers** | https://fep-fee.eu/ | 1967 | Belgium | Yes |
| **Frontiers** | https://www.frontiersin.org/ | 2007 | Switzerland | Yes |
| **Future Science Group** | http://www.future-science-group.com/ | 2001 | England | Yes |
| **Geological Society of London** | https://www.geolsoc.org.uk/publications | 1807 | England | Yes |
| **Geoscience Frontiers** | https://www.journals.elsevier.com/geoscience-frontiers | N/A | China | Yes |
| **Hapres** | https://www.hapres.com/ | N/A | N/A | No |
| **Henry Stewart Talks** | http://www.hstalks.com/ | 2003 | England | No |
| **Highwire** | https://www.highwirepress.com/ | 1995 | USA | No |
| **Hogrefe** | http://www.hogrefe.de/ | 1949 | Germany | No |
| **Horticultural Research** | https://academic.oup.com/hr | 1962 | USA | No |
| **ICSTI Int. Council for Scientific & Technical Information** | http://www.icsti.org/ | 1984 | France | No |
| **IEEE** | http://www.ieee.org/ | 1963 | USA | Yes |
| **Igaku-Shoin Ltd** | http://www.igaku-shoin.co.jp/ | 1944 | Japan | No |
| **Institution of Engineering and Technology** | https://www.theiet.org/ | 2006 | England | No |
| **IntechOpen** | https://www.intechopen.com/ | 2004 | England | Yes |
| **Inter-Research Science Publisher** | https://www.int-res.com/ | 1984 | Germany | No |
| **International Atomic Energy Agency** | http://www.iaea.org/ | 1957 | Austria | No |
| **International Commission on Illumination (CIE)** | http://www.cie.co.at/ | 1913 | Switzerland | No |
| **International Federation of Reproduction Rights Organisations** | http://www.ifrro.org/ | 1980 | Denmark | Yes |
| **International Publishers Association** | http://www.internationalpublishers.org/ | 1896 | France | No |
| **IOP Publishing** | http://www.ioppublishing.org/ | 1874 | England | Yes |
| **IP Innovative Publication Private Ltd** | https://www.ipinnovative.com/ | 2010 | India | No |
| **Ishiyaku Publishers Inc.** | https://www.ishiyaku.co.jp/index.aspx | 1921 | Japan | No |
| **ITHAKA S+R** | http://www.sr.ithaka.org/ | 2000 | USA | No |
| **IWA Publishing** | http://www.iwapublishing.com/ | 1998 | England | Yes |
| **Jenny Stanford Publishing** | https://www.jennystanford.com/ | N/A | N/A | No |
| **John Benjamins Publishing Company** | http://www.benjamins.nl/ | 1963 | Netherlands | Yes |
| **Johnson Matthey Technology Review** | https://technology.matthey.com/ | 1957 | England | No |
| **JOSPT** | https://www.jospt.org/ | 1979 | USA | No |
| **Karger Publishers** | http://www.karger.com/ | 1890 | Germany | Yes |
| **Ke Ai** | http://www.keaipublishing.com/ | 2013 | China | Yes |
| **Knowledge Futures Inc.** | https://www.knowledgefutures.org/ | 2018 | USA | No |
| **Kriyadocs** | https://www.kriyadocs.com/ | 2014 | India | No |
| **Kudos** | https://www.growkudos.com/ | 2012 | England | No |
| **Kugler Publications** | https://kugler.pub/ | 1974 | Netherlands | No |
| **LibLynx** | http://www.liblynx.com/ | 2014 | USA | No |
| **Mark Allen Group** | http://www.markallengroup.com/ | 1985 | England | No |
| **Materials Research Forum LLC** | http://www.mrforum.com/ | 2015 | USA | No |
| **Maverick** | https://www.maverick-os.com/ | 2008 | England | No |
| **McGraw-Hill Professional** | https://www.mhprofessional.com/ | 1966 | USA | No |
| **MDPI** | https://www.mdpi.com/ | 1996 | Switzerland | Yes |
| **Morgan and Claypool Publishers** | https://morganclaypoolpublishers.com/ | 2002 | USA | No |
| **Morressier** | https://www.morressier.com/ | 2014 | Germany | No |
| **Nankodo** | https://www.nankodo.co.jp/ | 1879 | Japan | No |
| **National Information Standards Organization** | http://www.niso.org/ | 1939 | USA | No |
| **New England Journal of Medicine** | https://www.nejm.org/ | 1811 | England | Yes |
| **Nova Techset** | https://novatechset.com/ | 1989 | India | No |
| **NUV - Nederlands Uitgeversverbond (Dutch Publishers Association)** | https://www.mediafederatie.nl/ | N/A | Netherlands | No |
| **OAE Publishing** | https://www.oaepublish.com/ | 2015 | USA | No |
| **Open Exploration Publishing Inc.** | https://www.explorationpub.com/ | 2019 | USA | No |
| **Optica Publishing Group** | https://opg.optica.org/ | 1916 | USA | Yes |
| **OSDEL - Greek Collecting Society for Literary Works** | https://www.osdel.gr/en/ | 1997 | Greece | No |
| **Oxford University Press** | https://corp.oup.com/ | 1586 | England | Yes |
| **Pharmaceutical Press** | https://www.pharmaceuticalpress.com/ | 1841 | England | No |
| **PHI Learning** | https://www.phindia.com/ | 1963 | India | No |
| **Portico** | https://www.portico.org/ | 2005 | USA | No |
| **Portland Press** | https://portlandpress.com/ | 1911 | England | No |
| **PSI** | https://www.psiregistry.org/ | N/A | England | No |
| **Publishers Association** | https://www.publishers.org.uk/ | 1896 | England | No |
| **Radcliffe Cardiology** | https://www.radcliffecardiology.com/ | 1987 | England | No |
| **Research Consulting** | https://www.research-consulting.com/ | 2013 | England | No |
| **Royal Society of Chemistry** | https://www.rsc.org/ | 1980 | England | Yes |
| **S. Hirzel Verlag** | https://www.hirzel.de/ | 1853 | Germany | No |
| **SAE International** | https://www.sae.org/ | 1905 | USA | Yes |
| **SAGE Publishing** | https://us.sagepub.com/en-us/nam/home | 1965 | USA | Yes |
| **Scion Publishing** | https://scionpublishing.com/ | 2003 | England | No |
| **SciPubLaw** | https://scipublawn.com/ | 2018 | USA | No |
| **Seismological Society of America** | https://www.seismosoc.org/ | 1906 | USA | No |
| **Silverchair** | https://www.silverchair.com/ | 1997 | USA | No |
| **SLACK Incorporated** | http://www.slackinc.com/ | 1962 | USA | Yes |
| **Society for Scholarly Publishing** | https://www.sspnet.org/ | 1978 | USA | No |
| **Springer Nature** | https://www.springernature.com/gp | 2015 | Germany | Yes |
| **Springer Publishing Company** | https://www.springerpub.com/ | 1950 | USA | No |
| **SPUR Infosolutions** | https://www.spurinfo.com/ | 2016 | India | No |
| **Straive** | https://www.straive.com/ | 1980 | Singapore | No |
| **Syndicat National de L'Edition (SNE)** | https://www.sne.fr/ | 1874 | France | No |
| **Taylor & Francis** | http://www.taylorandfrancis.com/ | 1852 | England | Yes |
| **The Chemical Society of Japan** | https://www.chemistry.or.jp/en/ | 1878 | Japan | No |
| **Thieme Publishing Group** | https://www.thieme.com/ | 1886 | USA | Yes |
| **Trans Tech Publications** | https://www.igpublish.com/trans-tech-publications/ | 1967 | Switzerland | No |
| **TrendMD** | https://www.trendmd.com/ | 2013 | Canada | No |
| **Tsinghua University Press** | http://www.tup.tsinghua.edu.cn/en/index.html | 1980 | China | No |
| **UNE - Spanish Association of University Presses** | https://www.unebook.es/es/ | 1987 | Spain | No |
| **Virtus Interpress** | https://virtusinterpress.org/ | 2003 | Ukraine | No |
| **VTex** | https://vtex.lt/ | 2011 | Lithuania | No |
| **W.W. Norton & Company** | https://wwnorton.com/ | 1923 | USA | No |
| **Wiley** | https://www.wiley.com/ | 1807 | USA | Yes |
| **Wolters Kluwer Health** | https://www.wolterskluwer.com/en-ca/health | 1978 | USA | No |
| **World Health Organization** | https://www.who.int/ | 1948 | Switzerland | No |
| **World Scientific Publishing** | https://www.worldscientific.com/ | 1981 | Singapore | Yes |
| **Xia & He Publishing Inc.** | https://www.xiahepublishing.com/ | 2011 | USA | Yes |
| **Xpublisher GmbH** | https://www.xpublisher.com/en | 2010 | Germany | No |
| **Total Policy Count** |  |  |  | **56** |

Of the 163 STM academic publishers, fifty-six (34.4%) provided guidance for authors on the use of generative AI technologies/chatbots in one or more aspects of preparing the manuscript (e.g. specific disclosure requirement, formal research methods, proofreading). Policies were extracted from separate publisher web pages, editorials, or brief statements embedded within the general author guidelines. Although some academic publishers had listed various aspects of their policies on different webpages, the publishers were not considered to have more than one policy (i.e., a maximum of one policy per publisher). The quantitative analysis of extracted data, reviewed for inconsistencies by a third contributor, are summarized in **Table 2**. Counts are also expressed as percentage values of the total number of STM academic publishers.

**Table 2.**
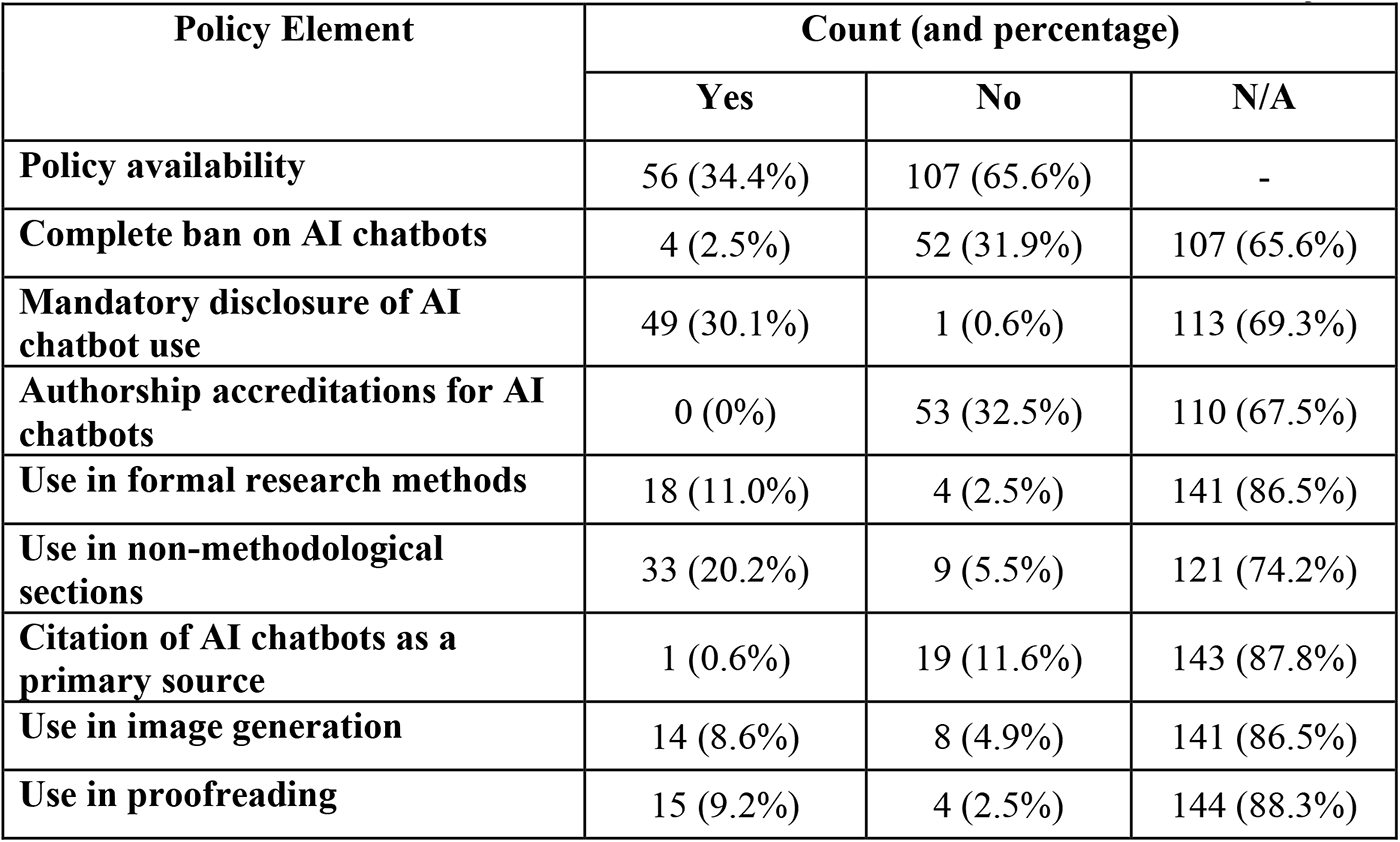
Data-Extracted Items of Academic Publisher Policies: A Summary.

Of the 56 academic publishers that had publicly available policies at the time of data collection, four (7.1%) (*AAAS, American Society of Civil Engineers, American Society of Mechanical Engineers,* and *EB Medicine*) stated a complete ban on the use of AI chatbots by authors. Additionally, nearly all (49/56 or 87.5%) policies mandated disclosure of the authors’ use of AI chatbots in either the ‘Methods’ or ‘Acknowledgements’ section of the manuscript. *Elsevier* and *Geoscience Frontiers* provided a specific disclosure statement to be used in a separate section of the manuscript. Only the *Federation of European Publishers* stated that the disclosure of AI chatbot use by authors may not be mandatory if the generative technology was used only as a “tool in the creation process” [16].

The majority of the policies (53/56 or 94.6%) stated that authorship may not be granted for AI chatbots (i.e., do not list the chatbot as an author). Three publisher policies did not have a specific statement for AI authorship, however, no academic publishers (0%) openly allowed AI chatbots to receive authorship. Only one academic publisher policy, *Karger Publishers*, stated that the AI technology used by authors must be cited as a primary source in the references. Nineteen (11.6%) publishers explicitly stated that AI chatbots should not be cited as primary sources.

Eighteen (11.0%) publishers allowed the use of AI chatbots in formal research methods, such as data organization and analysis, simulation and predictive modeling, and natural language processing. Most academic publisher policies generally stated, “formal research methods”, without specifying the range of permitted tasks. Additionally, 33/163 (20.2%) of academic publishers permitted the use of AI chatbots in non-methodological sections, including the process of writing the introduction and background sections. None of these 33 policies contained disclosure of any content check or plagiarism measures that the academic publisher or individual journal may implement to confirm the veracity of AI-generated work.

Finally, fourteen (8.6%) academic publishers permitted the use of AI technologies by authors to generate images; however, some policies also specified that significantly altering the properties of existing images is prohibited. Fifteen (9.2%) academic publishers explicitly granted authors the ability to use AI chatbots for proofreading the manuscript.

## Discussion

The objective of this study was to explore the approaches taken by scientific, technical, and medical publishers to regulate the use of AI chatbots by authors by examining their publicly available policies. There was no formal hypothesis regarding the approaches of academic publishers conveyed through their policies. This cross-sectional audit found that only about one-third of all STM academic publishers contained policies (56/163) on individual web pages or embedded in the author guidelines as of December 2023. Some academic publishers may have policies that are in development, individualized to journals within their portfolios, or not available for public view. Most policies required authors to disclose the use of AI chatbots in their manuscript submissions, usually for increased transparency and the contextual understanding of the need for AI chatbot use. Additionally, although most of the policies (33/56) permitted authors to use AI chatbots for methodological and non-methodological sections (e.g. assistance in writing the introduction), no academic publishers allowed AI chatbots to be listed as co-authors of manuscripts. For example, large and influential academic publishers, such as *Elsevier* and *Springer Nature*, stated that AI chatbots cannot receive authorship, although their use for formal research design protocols is permitted [17, 18]. This element-based analysis helped interpret the various approaches that academic publishers took to define the role of AI chatbots in medical research.

### Comparative Literature

Despite the novelty of the academic publisher policies introduced for the responsible use of AI chatbots by authors, there are similar cross-sectional audits examining academic publisher or journal policies for authors’ use of AI chatbots, as of April 2024 [5, 19–25]. There are also scoping reviews and meta-analyses examining the policies and attitudes of educational institutions, libraries, and other individual studies that explore the role of ChatGPT, particularly, in scientific and medical research [21–23]. However, only the former cross-sectional audits are considered directly relevant literature for comparison to this study.

For example, Lund and Naheem analyzed the policies of the top 300 academic journals, based on their SCImagoJR indexing factor, in late-spring 2023 [5]. They noted that over half (58.7%) of the examined journals had publicly available policies (176/300) to guide the use of AI generative technologies. Lund and Naheem also found that most of these guidelines were provided by the publisher rather than the individual journal [5]. Given that most were publisher-level policies, their content analysis showed similar findings to this study. For example, very few journals placed a ban on the authors’ use of AI chatbots (3.4%), almost all policies prohibited the accreditation of AI chatbots as authors (98.9%), and the majority required disclosure, primarily in the methods section (undisclosed percentage) [5].

Similarly, Ganjavi et al. examined the policies on the use of AI generative technologies for authors in the “top 100 academic publishers” and “top 100 highly ranked” (according to the SCImagoJR indexing factor) academic journals in summer 2023 [19]. Among these publishers and journals, the researchers found policies for 24% and 87%, respectively, with similar percentages reflecting that 96% and 98%, respectively, do not allow AI chatbots to be listed as authors. However, there was also some variability between the policies of some journals and their respective publisher [19].

Additionally, an analysis of the 25 largest journals in the fields of cardiology and cardiovascular medicine found that all permitted the documented use of AI chatbots by authors but did not require accreditation as a co-author or for the purposes of image generation [24]. These similar results suggest that large and influential academic publishers and journals, defined by the SCImago indexing factor, are in accordance with the necessary elements of their policy regulating authors’ use of AI chatbots, such as the listing of AI chatbots as co-authors.

### Implications and Future Directions

This cross-sectional audit suggests that some STM academic publishers have quickly responded in an attempt to protect the integrity and quality of their published content with policies that can guide the authors’ use of AI chatbots in research and publication. The results of this study provide insights into the approaches used by STM academic publishers to regulate AI use by authors whilst maintaining scientific rigour. These insights may inform other academic publishers, librarians, indexing services (e.g. PubMed, Web of Science), and the larger scientific community about how researchers may responsibly optimize AI chatbot use.

A common theme uncovered in this study and comparative literature is the restriction on listing AI tools as co-authors despite the permitted, declared use of the AI chatbot(s) [5, 19–25]. In fact, common ethics forums, such as the Committee of Publishing Ethics (COPE) and the International Committee of Medical Journal Editors (ICMJE), also propounded that this criterion to be adopted by academic publishers. The COPE declared that generative AI tools are non-legal entities, therefore, AI chatbots can neither take responsibility for the manuscript nor manage conflicts of interest [26]. Moreover, information generated by AI chatbots may be inaccurate, and prone to errors and biases [1, 22]. For instance, until recently, ChatGPT had not been updated on events and developments past January 2022, leading to unreliable information on current scientific findings [22]. Hence, these publisher policies may serve to inform researchers that authors, not AI chatbots, must take responsibility for the AI-generated content in their manuscript and would be held accountable for any content inaccuracies or breaches of publication ethics [26].

The regulations imposed by these policies may reflect the academic publishers’ consideration of the benefits and challenges of authors using AI chatbots. A cross-sectional survey by Ng et al. in 2023 revealed that researchers, although having expressed interest in the applications of AI chatbots in scientific research, received inadequate training in AI tool usage by their academic institutions [27]. In addition to the current limitations of AI chatbots (e.g., unverified content generation), researchers using AI tools without formal training may result in consequences of poor content quality and/or misinformation [27]. Academic publishers, being aware of these shortcomings, may have established policies as a measured response to safeguard the integrity of their publications; holding authors accountable for AI-generated content may also help researchers obtain a greater understanding of the potential impacts associated with the improper use of AI chatbots.

The present audit found that many academic publishers (i.e., 107/163 or 65.6% of STM academic publishers) do not yet have a policy regulating the use of AI chatbots by authors. Also, the 56 academic publishers that do have AI-chatbot policies for authors do not uniformly agree on key applications of AI chatbots, such as use in image generation and proofreading. Some policies may also have lacked clarity in certain aspects; for example, the range of permitted tasks for “formal research methods” was not specified. Future work may involve a re-examination of available policies of STM academic publishers (i.e., after 12-18 months) to uncover additional publisher policies that may currently be under construction and if existing policies have been updated to address some of the other applications of chatbots that were identified in this study (e.g. use in image generation, proofreading). Additionally, the use of AI chatbots by editors and peer-reviewers would also have important implications for the academic publishing industry, such as the fairness of editorial decisions on manuscripts [28]. Hence, a similar evaluation of the approaches or policies of academic publishers toward the use of AI chatbots by examining the policies for editors and peer-reviewers may be a next step of investigation. These inquiries would offer insights into the likely evolving ways that academic publishers establish the role of AI chatbots in the realm of research and publication.

### Strengths and Limitations

The STM association is the “leading globe trade association” for academic publishers (i.e., containing journals with the highest impact factors) that collectively publish roughly 66% of all peer-reviewed articles in the fields of science, technology, and medicine [15]. Therefore, understanding the nature of the policies of these academic publishers, thus far, may help in understanding their approaches and viewpoints about the use of AI chatbots by authors for manuscript submissions. The STM association provided a large sample of 56 of 163 academic publishers that had a policy, which should reflect in the range of approaches taken to regulate AI chatbot use by authors. Data extraction was performed independently and in duplicate by two authors, followed by data verification by a third contributor, helped ensure a consistent approach for the interpretation of academic publisher policies.

As this study provided a cross-sectional analysis of policies, it limits our understanding of the changes in approaches of academic publishers as compared to a longitudinal study. Most of the STM members are American or UK-based and this study did not examine non-English academic publishers or individual scholarly journals published in scientific communities, such as post-secondary institutions, that may vastly differ in attitudes toward the author usage of AI chatbots. Hence, the findings may not be generalizable to these excluded groups. We also did not assess the fidelity of publishers in following their own policies or the consistency of individual journals within the portfolio of each publisher.

## Conclusion

This audit examined the policies of academic publishers to regulate the use of AI chatbots by authors and found that only a third of STM academic publishers have posted policies. These policies showed considerable heterogeneity; many do not yet guide the use of AI for aspects such as image generation and formal research designs. Nevertheless, academic publishers find common ground in denying AI chatbots authorship accreditations, regardless of their level of contributions, which implies that authors are held accountable for the scientific rigour and integrity of the content produced by AI chatbots. The introduction of these policies suggests that many publishers are working to address the potential threats of the improper use of AI chatbots by authors. As the landscape of AI chatbots in research continues to evolve, academic publishers face the task of implementing appropriate and consistent measures to define and promote the responsible use of AI chatbots by authors.

## Data Availability

All relevant study materials and data are included in this manuscript or posted on the Open Science Framework.

https://doi.org/10.17605/OSF.IO/6HP9R

## List of Abbreviations

AI: artificial intelligence
ChatGPT: Chat Generative Pre-Trained Transformer
STM: Scientific, Technical, and Medical
OSF: Open Science Framework
COPE: Committee on Publishing Ethics
ICMJE: International Committee of Medical Journal Editors

## Declarations

### Ethics Approval and Consent to Participate

This article involved a review of publicly available publisher policies, and this did not require research ethics board approval.

### Consent for Publication

All authors consent to this manuscript’s publication.

### Availability of Data and Materials

All relevant study materials and data are included in this manuscript or posted on the Open Science Framework: <u>10.17605/OSF.IO/6HP9R</u>

### Authors’ Contributions

DB: collected and analysed data, co-drafted the manuscript, and gave final approval of the version to be published.

LD: collected and analysed data, co-drafted the manuscript, and gave final approval of the version to be published.

HJ: collected and analysed data, co-drafted the manuscript, and gave final approval of the version to be published.

CL: assisted with the design of the study and the analysis of data, made critical revisions to the manuscript, and gave final approval of the version to be published.

RBH: assisted with the design of the study and the analysis of data, made critical revisions to the manuscript, and gave final approval of the version to be published.

AI: assisted with the design of the study and the analysis of data, made critical revisions to the manuscript, and gave final approval of the version to be published.

AM: assisted with the design of the study and the analysis of data, made critical revisions to the manuscript, and gave final approval of the version to be published.

JYN: analyzed data, designed and conceptualized the study, made critical revisions to the manuscript and gave final approval of the version to be published.

